# Reliability, validity, and sensitivity of Japanese version of the UCLA Scleroderma Clinical Trial Consortium Gastrointestinal Tract Instrument: application to efficacy assessment of intravenous immunoglobulin administration

**DOI:** 10.1101/2023.09.19.23295773

**Authors:** Kazuki M Matsuda, Eiki Sugimoto, Yoshiaki Ako, Marie Kitamura, Mai Miyahara, Hirohito Kotani, Yuta Norimatsu, Teruyoshi Hisamoto, Ai Kuzumi, Takemichi Fukasawa, Shinichi Sato, Ayumi Yoshizaki

## Abstract

**Objective:** This study aimed to develop and assess the reliability, validity, and sensitivity of Japanese version of the University of California Los Angeles Scleroderma Clinical Trial Consortium Gastrointestinal Tract (GIT) Instrument 2.0 (the GIT score), as an evaluation tool for GIT symptoms in systemic sclerosis (SSc).

**Methods:** Japanese version of the GIT score was constructed using the forward-backward method. The reliability and validity of this instrument were evaluated in a cohort of 38 SSc patients. Correlation analysis was conducted to assess the relationship between the GIT score and existing patient-reported outcome measures. Additionally, the sensitivity of the GIT score was examined by comparing GIT scores before and after intravenous immunoglobulin (IVIG) administration in 10 SSc-myositis overlap patients, as IVIG has recently demonstrated effectiveness in alleviating GIT symptoms of SSc.

**Results:** Japanese version of the GIT score exhibited internal consistency and a significant association with the Frequency Scale for the Symptoms of Gastroesophageal Reflux Disease. Furthermore, the total GIT score, as well as the reflux and distention/bloating subscales, displayed moderate correlations with the EQ-5D pain/discomfort subscale, Short Form-36 body pain subscale, and its physical component summary. Notably, following IVIG treatment, there was a statistically significant reduction in the total GIT score and most of the subscales.

**Conclusion:** We firstly validated Japanese version of the GIT score in Japanese SSc patients in real-world clinical settings. This instrument holds promise for application in future clinical trials involving this patient population.

**Key messages:**

- What is already known about this subject? Khanna et al. developed the UCLA Scleroderma Clinical Trial Consortium Gastrointestinal Tract (GIT) Instrument (the GIT score) to assess patient-reported GIT symptoms in individuals with systemic sclerosis (SSc).
- What does this study add? We have developed and established the reliability, validity, and sensitivity of the Japanese version of the GIT score in cohorts of Japanese individuals with SSc.
- How might this impact on clinical practice? This tool can effectively evaluate GIT manifestations in Japanese SSc patients in routine clinical settings, and potentially in clinical trial contexts.

## Introduction

Systemic sclerosis (SSc) is a complex connective tissue disease typified by widespread inflammation, vasculopathy, and severe fibrosis affecting various organs including the skin, lungs, and the gastrointestinal tract (GIT).[1] The fibrotic process particularly compromises the GIT by inducing hypomotility, leading to a spectrum of manifestations throughout both the upper and lower GIT, such as gastroesophageal reflux disease (GERD) and intestinal pseudo-obstruction (IPO). Notably, as many as 90% of SSc patients suffer GIT abnormalities, which significantly associates with a marked decline in health-related quality of life (HRQOL),[2] extended duration of hospitalization, and in severe cases, increased mortality rates.[3]

Although the current therapeutic modalities for addressing the GIT involvement of SSc have been constrained, the emergence of innovative treatment approaches with disease-modifying potential, including biologics[4][5] and autologous hematopoietic stem cell transplantation,[6] harbors promise for more efficacious outcomes. Furthermore, recent investigations have underscored the advantages of intravenous immunoglobulin (IVIG), one of the conventional agents being tried for SSc management characterized by its low adverse event profile, for mitigating the GIT symptoms of SSc.[7] As such, the development of clinical outcome measures that are robust, valid, and sufficiently sensitive for use in clinical trials is imperative to assess the efficacy of these groundbreaking therapies on the GIT symptoms of SSc.

The recent trend towards the integration of solid methodologies for capturing patient perspectives in clinical trials has assumed escalating importance in regulatory decision-making, aiming to enhance the ’patient-centeredness’ of drug development processes. Consequently, patient-reported outcome measures (PROMs) have become increasingly pertinent in the context of SSc. Given the substantial heterogeneity and complexity of the clinical manifestations in SSc patients, their evaluation necessitates a multidimensional approach. For instance, certain patients may exhibit severe symptoms related to upper GIT involvement, such as reflux, while others may predominantly present complaints attributed to lower GIT abnormalities, such as distention and bloating. Furthermore, conditions that appear mutually exclusive, such as diarrhea and constipation, may paradoxically coexist at varying times throughout the disease progression. This co-occurrence further complicates the comprehensive communication of the full spectrum of GIT symptoms between patients and clinicians.

In an effort to develop a PROM to subjectively and holistically quantify GIT involvement in SSc patients, Khanna et al. conceptualized the SSc-GIT 1.0 in 2007.[8] This tool was initially formulated as a 52-item questionnaire, the content of which was guided by an extensive literature review, expert consensus, and the findings from two focus groups. Subsequently, in 2009, Khanna et al. introduced a more concise and refined version known as the University of California Los Angeles (UCLA) Scleroderma Clinical Trial Consortium (SCTC) GIT 2.0 Instrument (the GIT score), which comprised of 34 items.[9] Evidence suggests that the GIT score exhibits commendable test-retest reliability. Further, both the total and subscale scores were shown to effectively differentiate between patients with mild, moderate, and severe self-rated GIT involvement. Thus, its application in both clinical trials and routine patient care has been strongly endorsed.

The GIT score has been adapted and validated in multiple languages, including but not limited to French,[10] Dutch,[11] Italian,[12] Romanian,[13] and Chinese.[14] Although a Japanese translation of the questionnaire has been made available by Khanna et al. online, it has yet to undergo validation within the Japanese population. In light of this, we undertook the reformation of the Japanese version of the GIT score, based on its original counterpart. This newly adapted tool was then implemented in a cohort of Japanese patients with SSc in our clinic, and its reliability and validity were evaluated using statistical methodologies. We also assessed the correlation between the GIT scores and clinical manifestations or autoantibody profiles of SSc patients in Japan.

Furthermore, we gauged the sensitivity of Japanese version of the GIT score by comparing scores before and after the administration of IVIG, which demonstrated rapid improvement of GIT symptoms of SSc in a previous study.[7] In this study, our primary objective is to establish this questionnaire as a benchmark tool for evaluating therapeutic efficacy in clinical trials involving the Japanese population.

## Materials and Methods

### Translation

We utilized the "forward-backward method"[15] to construct a Japanese adaptation of the GIT score. The process began with independent translations by two translators (KMM and ES), both native speakers of Japanese. They then came together to scrutinize each item, identifying and resolving any potential points of confusion or ambiguity until they reached a consensus. This intermediate version of the instrument was then tested on 10 non-bilingual SSc subjects with no issues arising in relation to clarity or comprehension. Subsequently, this version underwent back-translation by two bilingual translators. The English rendition produced from this process was critically reviewed by 2 native English speakers, who found no need for further modifications (**Supplementary Data 1**).

### Patients

We consecutively recruited Japanese patients with SSc visiting our scleroderma center outpatient clinic from November 2022 until April 2023 for assessing the reliability and validity of Japanese version of the GIT score. All the SSc patients fulfilled the classification criteria established by the American College of Rheumatology and European League Against Rheumatism in 2013.[16] We also sequentially enrolled patients with SSc-myositis overlap admitted to our wards from April 2023 until August 2023 for IVIG administration for evaluating sensitivity of Japanese version of the GIT score. This study was approved by The University of Tokyo Ethical Committee (Approval Number 0695). Written informed consent was obtained from all the human subjects.

### Clinical data acquisition

Clinical data were collected by retrospective review of electric medical records. We gathered basic patient information, symptoms, medications, and laboratory findings from the closest time point from the date of the GIT score evaluation. SSc patients were categorized by LeRoy’s classification rule into diffuse cutaneous SSc (dcSSc), limited cutaneous SSc (lcSSc), or overlap syndrome.[17] Skin thickness was semi-quantitatively examined by the modified Rodnan total skin thickness score (mRSS).[18] Interstitial lung disease (ILD), pulmonary hypertension (PH), and scleroderma renal crisis (SRC) were diagnosed as previously described.[19]

### Autoantibody detection

Autoantibodies in the serum samples were evaluated utilizing autoantibody array assay (A-Cube) as previously described.[20][21] Briefly, a total of 65 antigens of 43 autoantibodies associated with SSc, Sjogren syndrome (SjS), primary biliary cholangitis (PBC), myositis, and overlap syndrome, with FLAG-GST-tag on the N-terminus were synthesized *in vitro* with a wheat germ cell-free translation system,[22] from human cDNA library entry clones.[23] The synthesized proteins were captured on array plates under wet conditions by affinity between the GST tags and glutathione (GSH) coated over the glass slides.[24] Then the slides were consequently treated with serum samples diluted in the blocking buffer and fluorescence-labeled anti-human IgG antibody (Ab). After the slides were washed and air-dried, the plates were scanned by a fluorescence imager (**Supplementary Figure A**). The negative control spots were prepared using distilled water instead of mRNA during protein preparation. The positive control spots were prepared using mRNA encoding human IgG for protein synthesis. The autoantibody quantification was performed based on the fluorescent values obtained from reactions of serum with the protein spots. The level of each autoantibody was calculated as below:

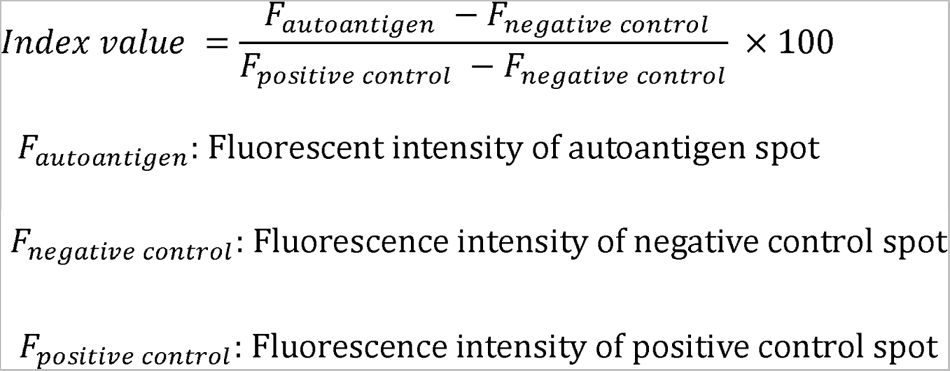

The cut-off value of each autoantigen was determined based on the mean + 3 standard deviation (SD) of healthy controls.

### Cytokine measurement

The serum levels of cytokines were measured by Luminex Discovery Assay Human Premixed Multi-Analyte Kit (R&D Systems, Minneapolis, MN, USA) according to the manufacturer’s protocol. The evaluated cytokines were as follows: tumor necrosis factor-alpha (TNF-α), interleukin 6 (IL-6), interleukin 10 (IL-10), interleukin 27 (IL-27), vascular endothelial growth factor (VEGF), interferon-gamma (IFN-γ), interleukin 31 (IL-31), interleukin 1 alpha (IL-1α), interleukin 4 (IL-4), interleukin 17 (IL-17), B cell activating factor belonging to the tumor necrosis factor family (BAFF), interleukin 13 (IL-13), interferon alpha (IFN-α), and interleukin 23 (IL-23).

### Patient-reported outcome measures

Patients completed the Japanese version of the GIT score, Medical Outcomes Short Form (SF)-36,[25] the EQ-5D with five levels tool,[26] and the F-scale.[27] UCLA SCTC GIT 2.0 comprises 34 items divided into seven domains: reflux, distention/bloating, diarrhea, fecal soilage, constipation, emotional well-being, and social functioning.[9] Each domain is rated from 0 (indicating better HRQOL) to 3 (representing poorer HRQOL), with the exception of the diarrhea and constipation domains, which have a range of 0–2 and 0–2.5, respectively. The overall GIT score is the mean score of six out of the seven domains, excluding constipation, and varies from 0 (higher HRQOL) to 3 (lower HRQOL). The original version in English is accessible online at http://uclascleroderma.researchcore.org/.

The SF-36 is a broad-spectrum measure of health status, comprised of 36 items that evaluate 8 distinct domains.[25] Four scales examine physical health, namely physical functioning (10 items), bodily pain (2 items), role limitations resulting from physical health perceptions (4 items), and overall health perceptions (5 items). An additional four scales are dedicated to mental health, which include mental health (5 items), role limitations due to emotional concerns (3 items), vitality (4 items), and social functioning (2 items), alongside a single-item health transition scale. The physical health scales collectively form the Physical Component Summary (PCS), and the mental health scales together make up the Mental Component Summary (MCS). These summarized scores are normalized to the general population in Japan, which is characterized by a mean ± SD score of 50 ± 10.[28] A standard 4-week recall period was implemented.

The EQ-5D questionnaire with five levels is a generic instrument to quantify HRQOL.[26] The EuroQoL Group developed and tested this tool for the purpose of providing measurable health outcomes. In an initial study with SSc patients, the Italian version of this tool proved to be valid.[12] The EQ-5D is composed of two primary components: the first section, known as the EQ-5D profile, generates a health profile derived from a descriptive system. This system defines health based on five dimensions: ’mobility’, ’self-care’, ’usual activities’, ’pain or discomfort’, and ’anxiety or depression’. Each dimension offers three response categories indicating no problems, some problems, or extreme problems. The second component of the questionnaire is the EQ-5D Visual Analogue Scale (VAS), which evaluates the overall HRQOL on a scale from 0 (the worst possible health state) to 100 (the best possible health state). For this study, a standard 4-week recall period was employed.

F-scale refers to the Frequency Scale for the Symptoms of GERD (FSSG), which is a self-report questionnaire used to assess the frequency and severity of GERD-related symptoms, originally developed in Japan.[27] The FSSG consists of 12 items grouped into two subscales: a reflux-related subscale (acid regurgitation and heartburn) and a dysmotility-related subscale (including symptoms like non-cardiac chest pain, a sensation of a lump in the throat, belching, etc.). Each item is rated on a 4-point scale (ranging from never = 0, occasionally = 1, sometimes = 2, often = 3), and the scores are added together to provide a measure of the severity of GERD symptoms.

### Statistical analysis

We analyzed average scores, SDs, ranges, and the percentage of missing data. The floor and ceiling effects of the GIT scorewere determined by calculating the percentage of participants who scored at the extreme lower (floor) and upper (ceiling) limits. We gauged the internal consistency of the GIT score through Cronbach’s alpha.[29] We evaluated the construct convergent validity by examining the relationship among the GIT score, the EQ-5D, and the SF-36 domains, using Spearman’s rho to measure correlations. The association between the GIT scores and clinical manifestations or autoantibody profiles was investigated by logistic regression analyses. Data analysis was performed using Stata 15/IC (StataCorp, College Station, TX, USA), GraphPad Prism 9 (GraphPad Software, Boston, MA, USA), R, RStudio, and R packages “dplyr”, ”ggplot2”, “hrbrthemes”, “ggcorrplot”, and “ComplexUpset”. We set the threshold for statistical significance at *P* < 0.05.

## Results

### Study population

We recruited 38 patients with SSc for the assessment of reliability and validity of Japanese SCTC UCLA GIT 2.0 (**Table 1**). A large majority were female (94%) with the mean age of 65 years with SD of 11 years, all of whom were of Japanese ethnicity. The proportion of the patients classified into dcSSc was 21%. None of the subjects within this cohort had been classified as overlap with myositis. Comprehensive autoantibody screening utilizing A-Cube revealed anti-centromere Ab, anti-topoisomerase I Ab, anti-RNA polymerase III Ab, and anti-U1-RNP Ab with the prevalence rate of 45%, 26%, 16%, and 5%, respectively (**Supplementary** Figure 1B and 1C).

**Table 1.**
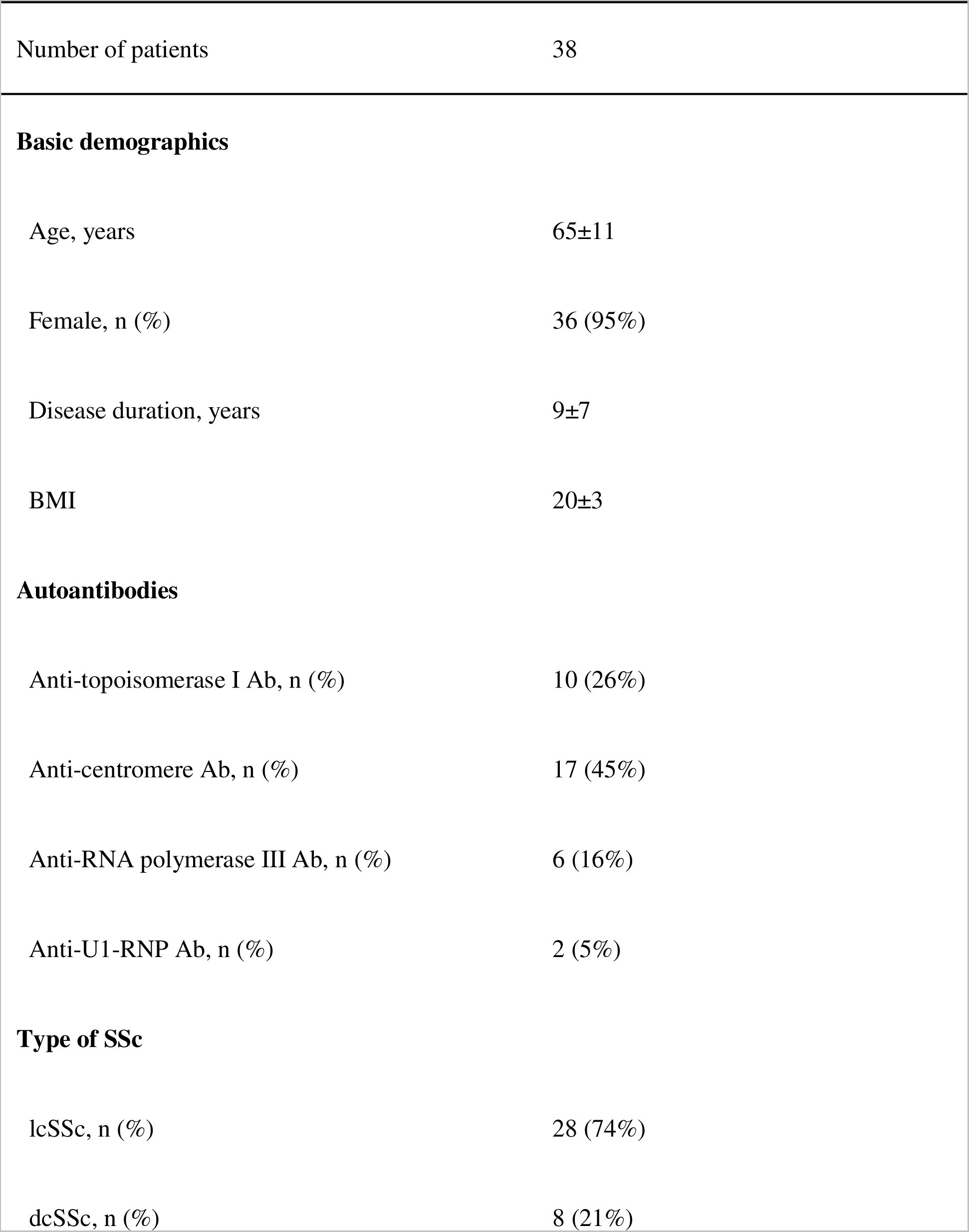
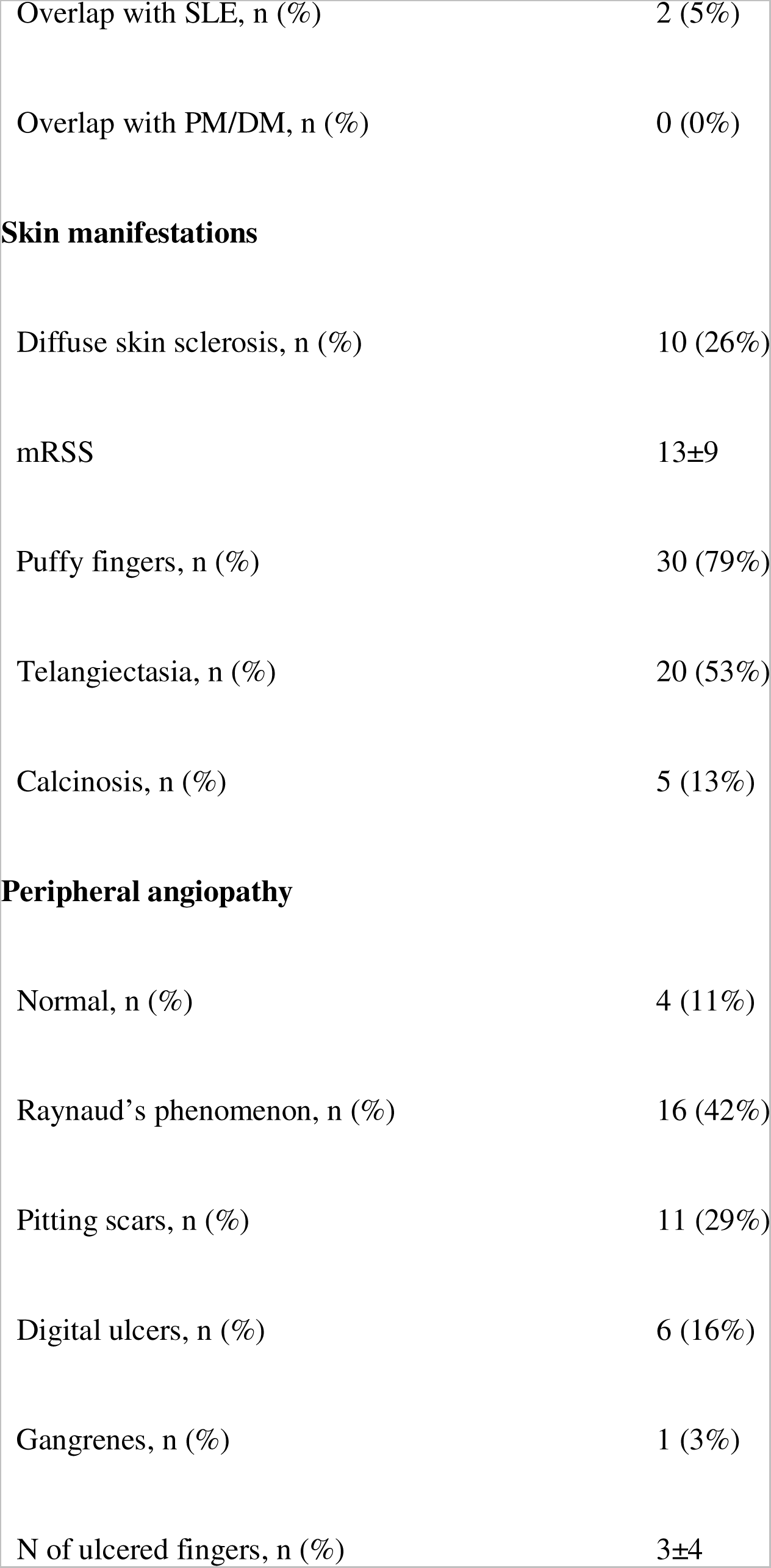
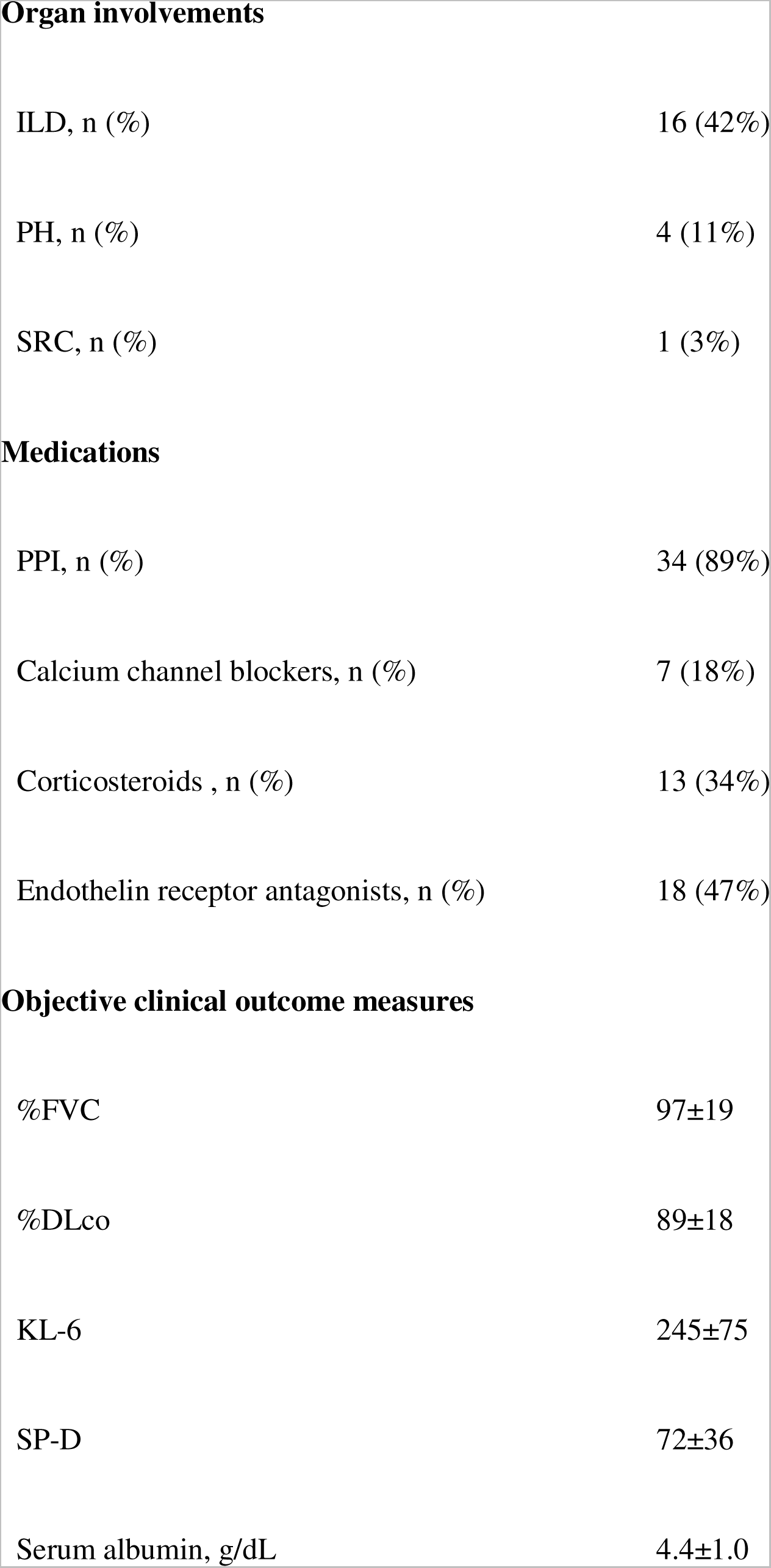
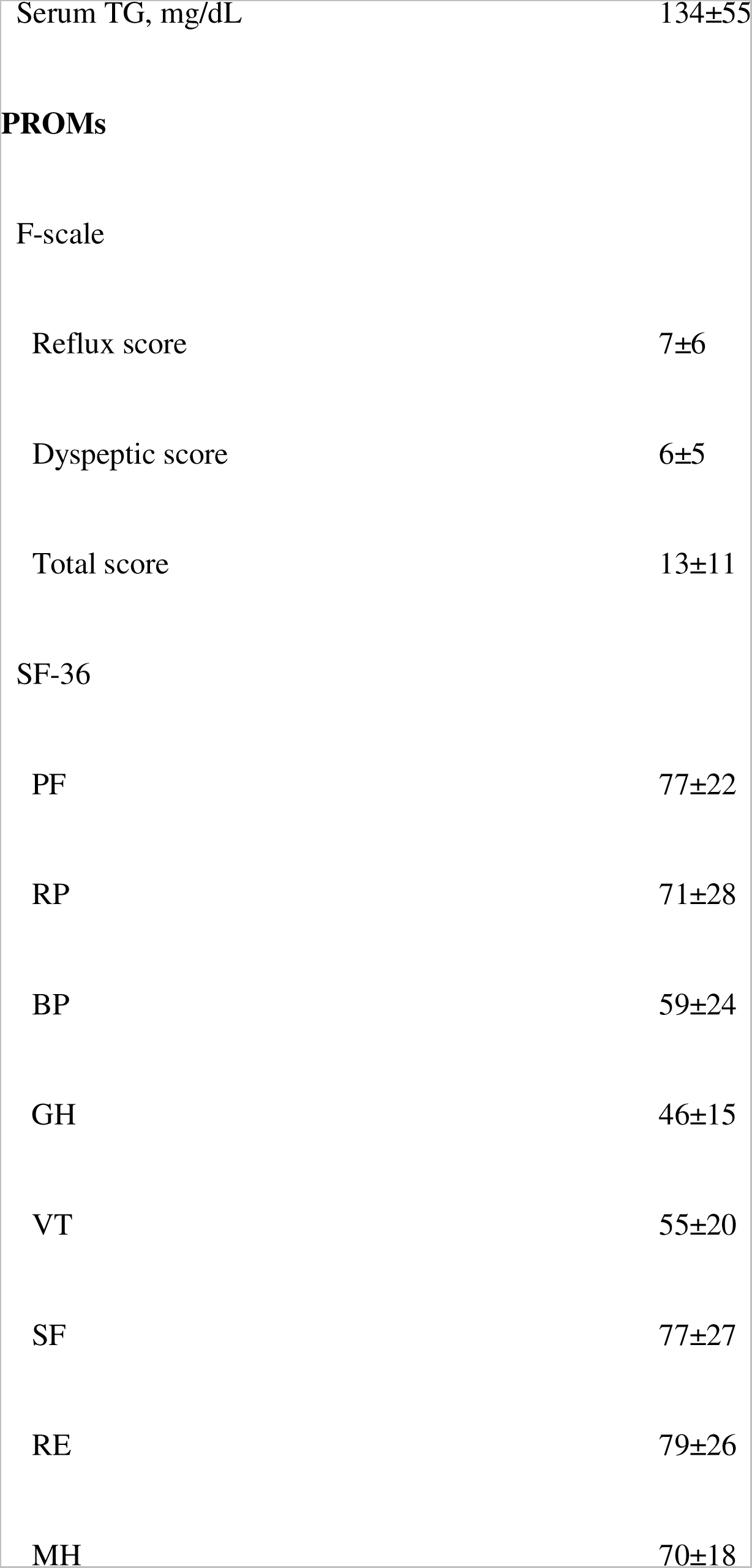
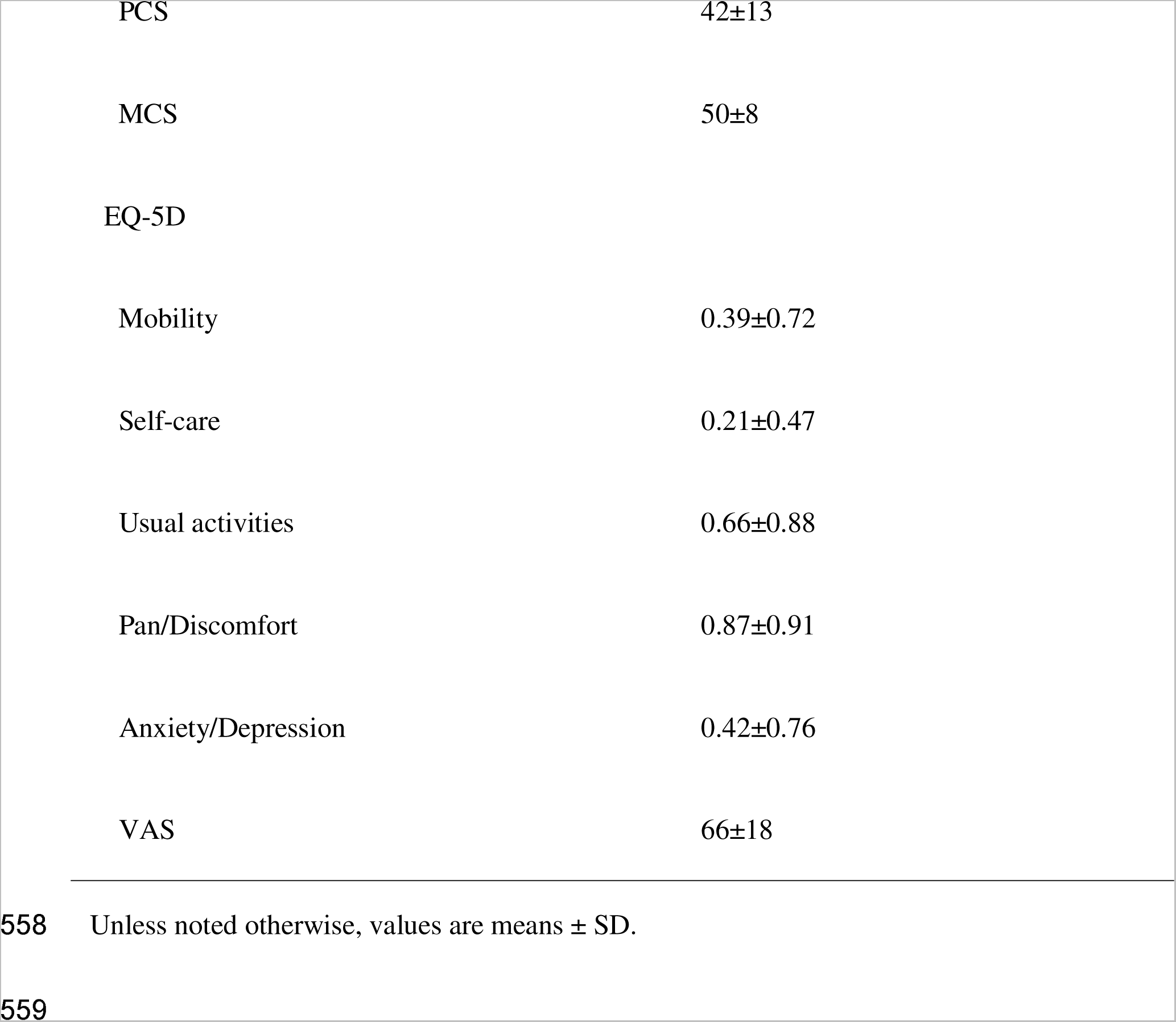
Background of the patients for reliability and validity assessment.

|  |  |
| --- | --- |
| Number of patients | 38 |
| **Basic demographics** |  |
| Age, years | 65±11 |
| Female, n (%) | 36 (95%) |
| Disease duration, years | 9±7 |
| BMI | 20±3 |
| **Autoantibodies** |  |
| Anti-topoisomerase I Ab, n (%) | 10 (26%) |
| Anti-centromere Ab, n (%) | 17 (45%) |
| Anti-RNA polymerase III Ab, n (%) | 6 (16%) |
| Anti-U1-RNP Ab, n (%) | 2 (5%) |
| **Type of SSc** |  |
| lcSSc, n (%) | 28 (74%) |
| dcSSc, n (%) | 8 (21%) |
| Overlap with SLE, n (%) | 2 (5%) |
| Overlap with PM/DM, n (%) | 0 (0%) |
| **Skin manifestations** |  |
| Diffuse skin sclerosis, n (%) | 10 (26%) |
| mRSS | 13±9 |
| Puffy fingers, n (%) | 30 (79%) |
| Telangiectasia, n (%) | 20 (53%) |
| Calcinosis, n (%) | 5 (13%) |
| **Peripheral angiopathy** |  |
| Normal, n (%) | 4 (11%) |
| Raynaud's phenomenon, n (%) | 16 (42%) |
| Pitting scars, n (%) | 11 (29%) |
| Digital ulcers, n (%) | 6 (16%) |
| Gangrenes, n (%) | 1 (3%) |
| N of ulcerated fingers, n (%) | 3±4 |
| **Organ involvements** |  |
| ILD, n (%) | 16 (42%) |
| PH, n (%) | 4 (11%) |
| SRC, n (%) | 1 (3%) |
| **Medications** |  |
| PPI, n (%) | 34 (89%) |
| Calcium channel blockers, n (%) | 7 (18%) |
| Corticosteroids , n (%) | 13 (34%) |
| Endothelin receptor antagonists, n (%) | 18 (47%) |
| **Objective clinical outcome measures** |  |
| %FVC | 97±19 |
| %DLco | 89±18 |
| KL-6 | 245±75 |
| SP-D | 72±36 |
| Serum albumin, g/dL | 4.4±1.0 |
| Serum TG, mg/dL | 134±55 |
| Reflux score | 7±6 |
| Dyspeptic score | 6±5 |
| Total score | 13±11 |
| SF-36 |  |
| PF | 77±22 |
| RP | 71±28 |
| BP | 59±24 |
| GH | 46±15 |
| VT | 55±20 |
| SF | 77±27 |
| RE | 79±26 |
| MH | 70±18 |
| PCS | 42±13 |
| MCS | 50±8 |
| EQ-5D |  |
| Mobility | 0.39±0.72 |
| Self-care | 0.21±0.47 |
| Usual activities | 0.66±0.88 |
| Pain/Discomfort | 0.87±0.91 |
| Anxiety/Depression | 0.42±0.76 |
| VAS | 66±18 |

Unless noted otherwise, values are means ± SD.

### Reliability

The average GIT score was 0.29 (SD 0.33), with 24% reporting no symptoms (total score = 0), 55% reporting mild symptoms (total score = 0.01–0.49), 16% reporting moderate symptoms (total score = 0.50–1.00), and 5% reporting severe symptoms (total score = 1.01–3.00). All multi-item subscales displayed a Cronbach’s alpha ranging from 0.67 to 0.92 (**Table 2**). A significant floor effect was evident for the total score and all its subscales, ranging from 24% (total score) to 89% (fecal soilage), while there was no observable ceiling effect.

**Table 2.** Descriptive statistics and internal consistency reliability of the Japanese.

| Subclass | Mean<br>(SD) | Min | Max | Cronbach's<br>$\alpha$ | Floor<br>effect<br>(%) | Ceiling<br>effect<br>(%) |
| --- | --- | --- | --- | --- | --- | --- |
| Reflux | 0.25<br>(0.37) | 0 | 1.625 | 0.77 | 17<br>(45%) | 0 (0%) |
| Distention/bloating | 0.70<br>(0.67) | 0 | 2.25 | 0.76 | 12<br>(32%) | 0 (0%) |
| Fecal soilage | 0.13<br>(0.41) | 0 | 2 | NA | 34<br>(89%) | 0 (0%) |
| Diarrhea | 0.49<br>(0.68) | 0 | 3 | 0.67 | 20<br>(53%) | 1 (%) |
| Social functioning | 0.21<br>(0.37) | 0 | 1.5 | 0.76 | 24<br>(63%) | 0 (0%) |
| Emotional<br>well-being | 0.17<br>(0.33) | 0 | 1.67 | 0.78 | 21<br>(55%) | 0 (0%) |
| Constipation | 0.36<br>(0.53) | 0 | 1.75 | 0.73 | 19<br>(50%) | 0 (0%) |
| Total GIT score | 0.29<br>(0.33) | 0 | 1.27 | 0.92 | 9<br>(24%) | 0 (0%) |

NA: not applicable.

### Validity

The reflux subscale and the distention/bloating subscale of the GIT score showed strong and significant correlation with the total score, the reflux subscale, and the dyspepsia subscale of the F-scale (**Table 3**). The total GIT score, the reflux and distention/bloating subscales also demonstrated moderate correlations with the EQ-5D pain/discomfort subscale, the SF-36 BP subscale, and the SF-36 physical component summary. Furthermore, there was a statistically significant, although weak, correlation between selected GIT subscales and certain SF-36 domains and components.

**Table 3.**
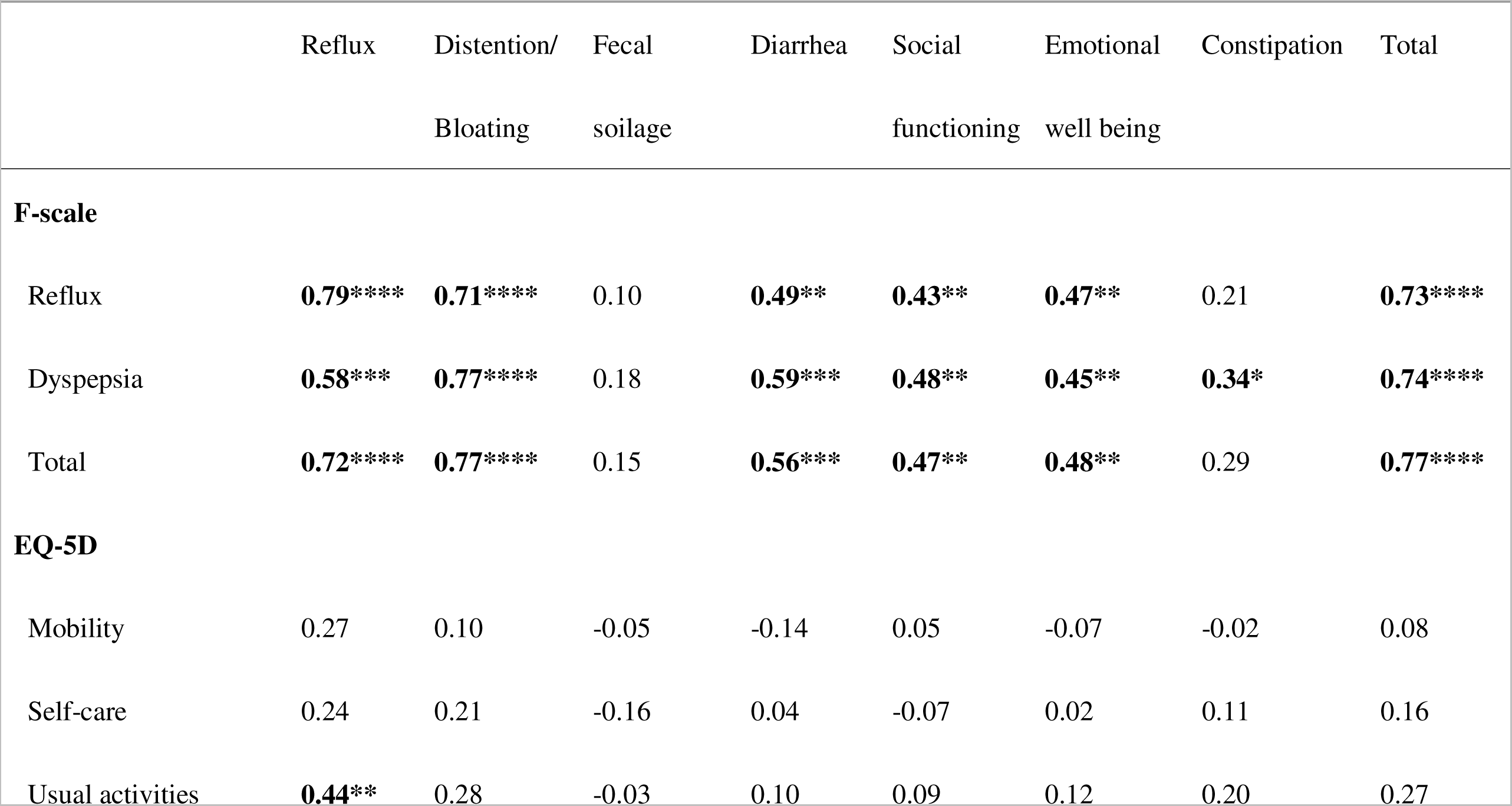
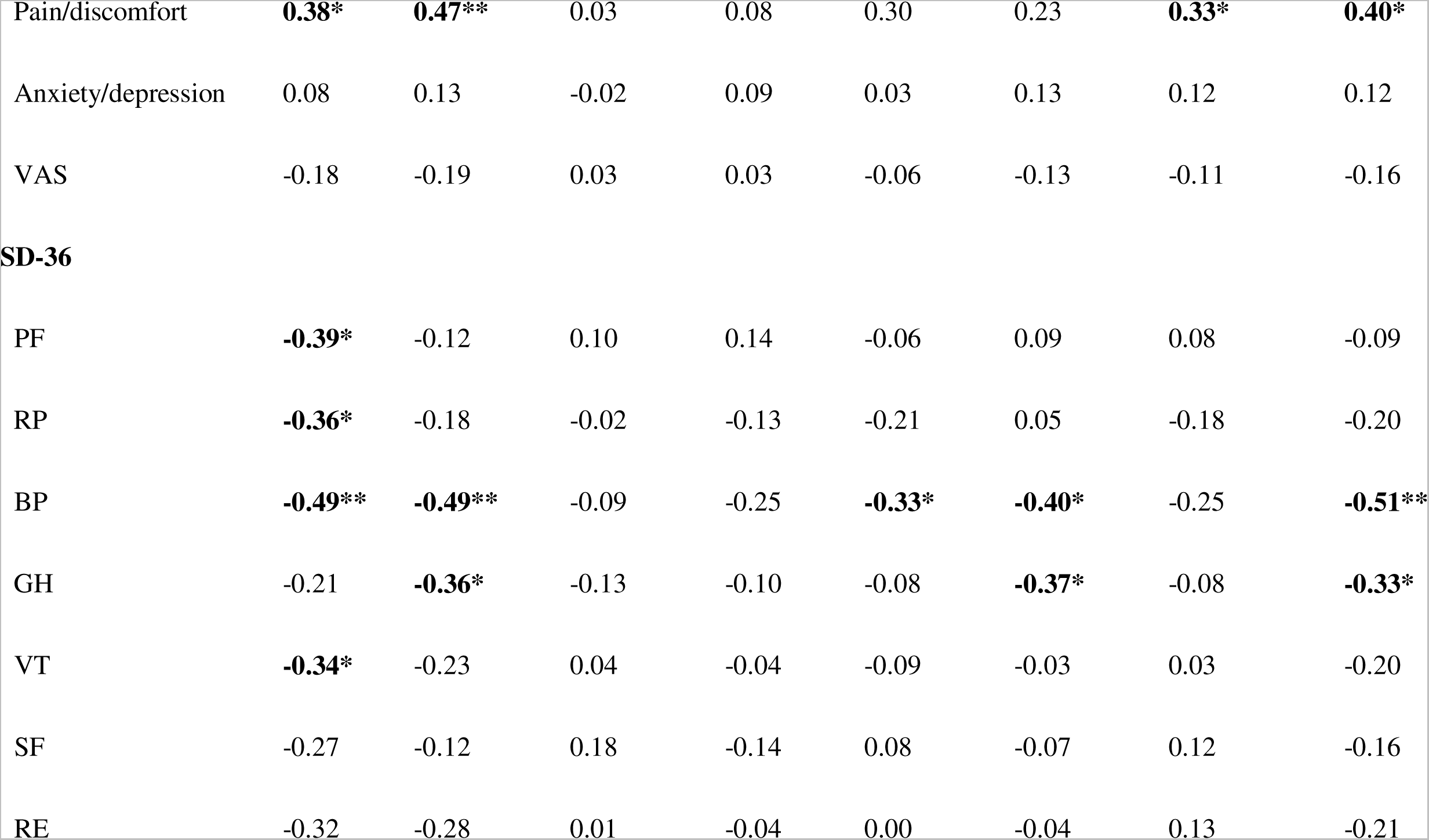
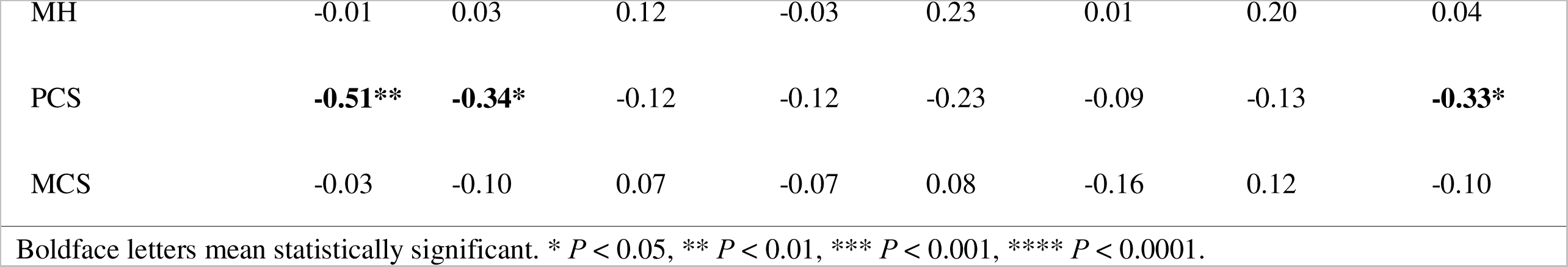
Spearman’s correlation coefficients among PROMs.

### Association between clinical features

No statistically significant correlation was observed between the GIT scores and the clinical manifestations of SSc, as indicated in **Supplementary Table 1**. Similarly, there was no significant association between the GIT scores and Ab profiles, as detailed in **Supplementary Table 2**. Meanwhile, a statistically significant correlation was observed between the serum levels of several cytokines and specific GIT subscales, as outlined in **Table 4**. Notably, there was a significant correlation between serum levels of TNF-α or IL-6 and the reflux subscale, as illustrated in **Figure 1A and 1B**. Additionally, a significant correlation was found between serum levels of VEGF and the social functioning or constipation subscales, as depicted in **Figure 1C** and **1D**.

**Figure 1.**
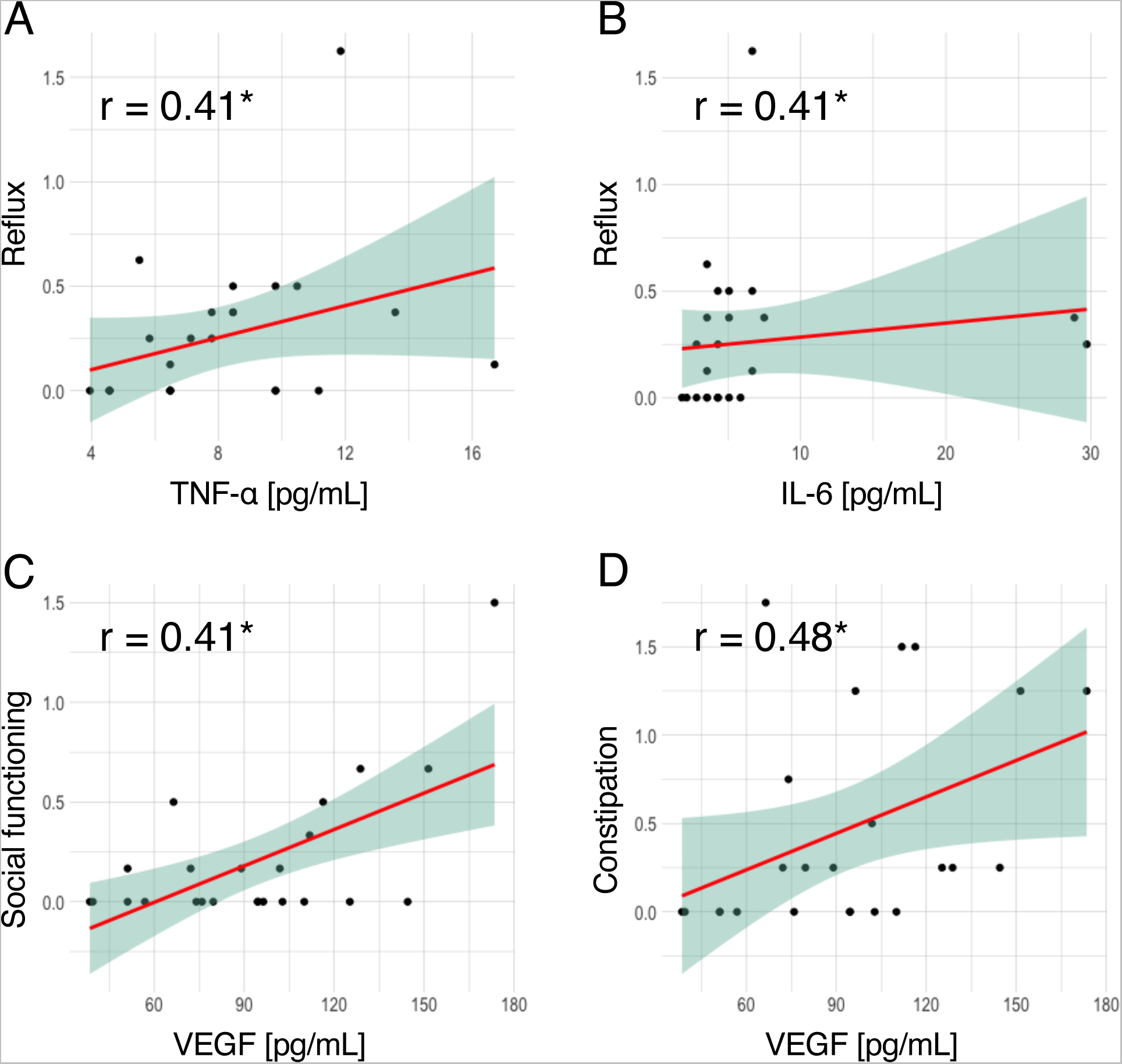
Correlation between serum cytokine levels and Japanese UCLA SCTC GIT 2.0 scores. Scatter plots of TNF-α vs. reflux subscale (**A**), IL-6 vs. reflux subscale (**B**), VEGF vs. social functioning subscale (**C**), and VEGF vs. constipation subscale (**D**). r: Spearman’s rho. * *P* < 0.05. The red line and the green area represent the regression line and its 95% confidence interval.

**Table 4.**
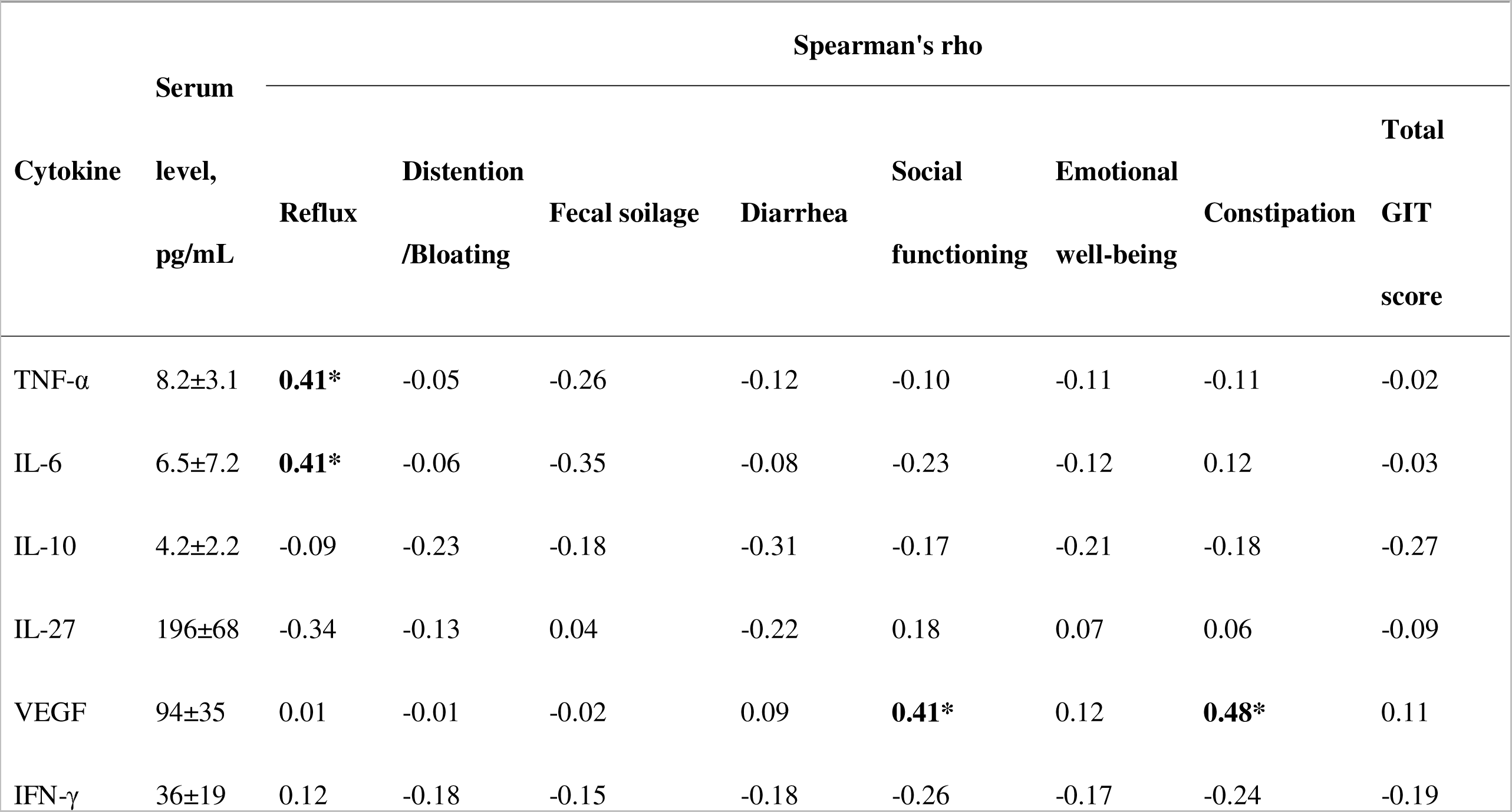
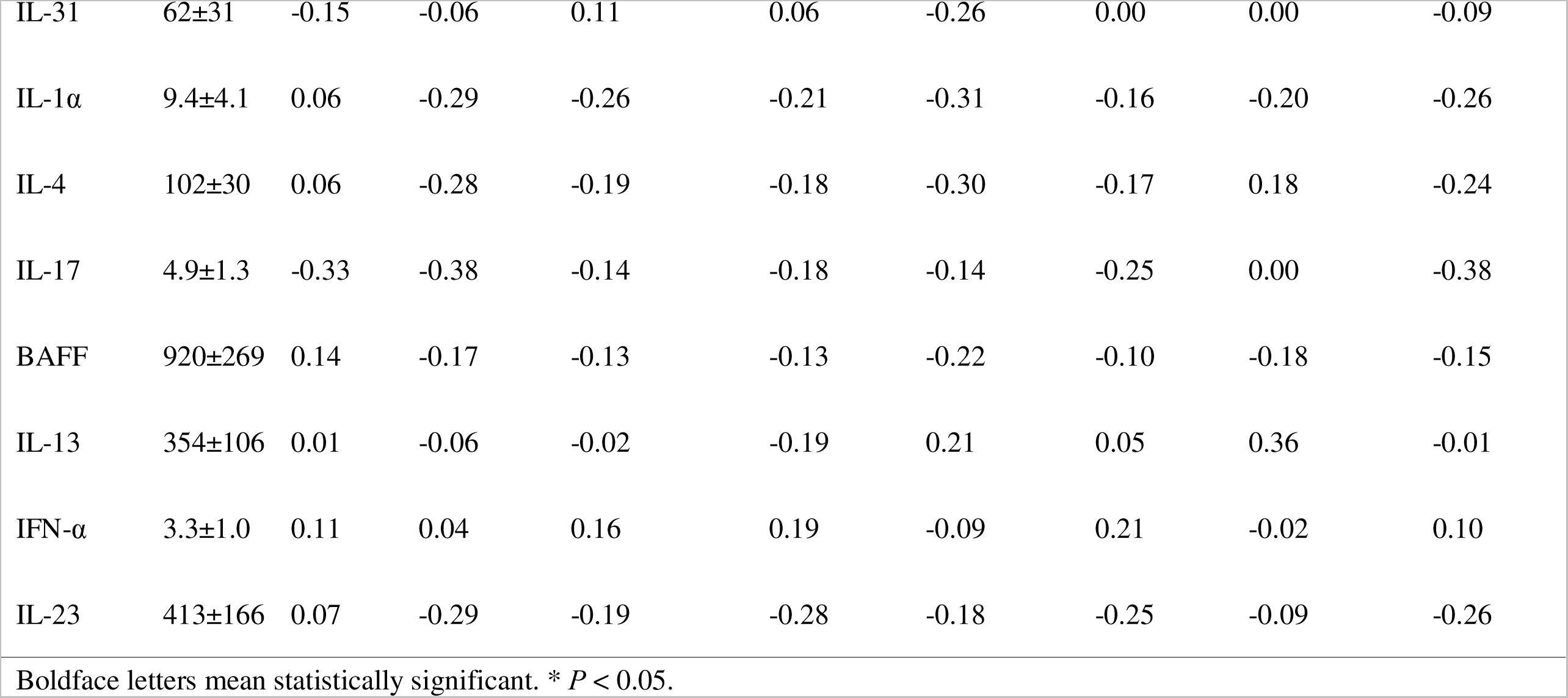
Association between serum cytokine levels and the UCLA SCTC GIT scores.

### Sensitivity

We enrolled a cohort of 10 Japanese patients diagnosed with SSc-myositis overlap, with a predominance of 9 female patients (90%). Their average age was 65 years, with a SD of 8 years. Among the patients, 6 patients were positive for anti-centromere Ab, 2 patients were positive for anti-U3-RNP Ab positivity, and one patient was positive for anti-RNA polymerase III Ab. Japanese version of the GIT score was administered both before and after IVIG treatment (**Figure 2A**), revealing a reduction in total GIT scores with statistical significance, as well as in a large part of the subscales, except for fecal soilage and constipation (**Figure 2B**).

**Figure 2.**
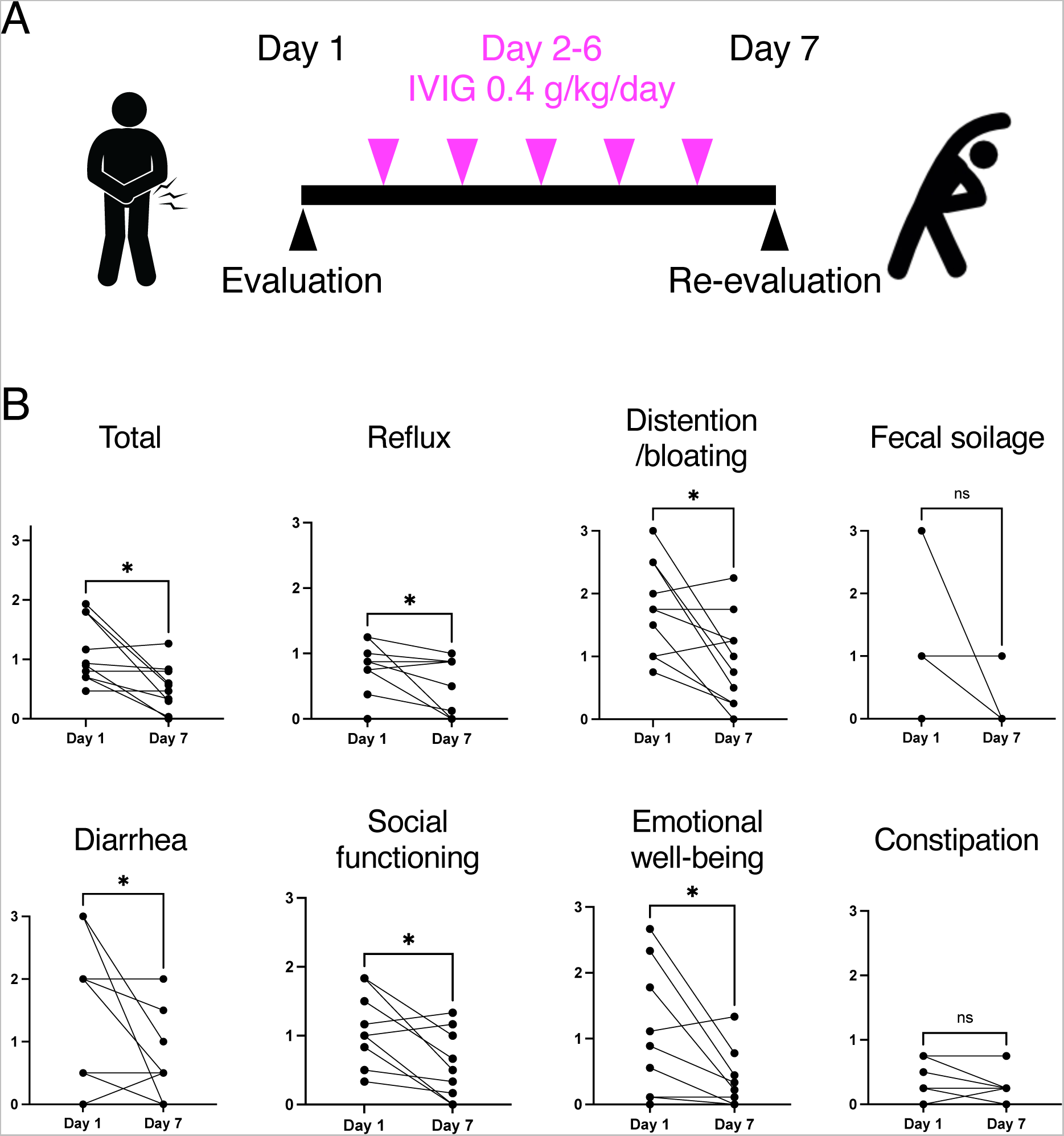
Sensitivity of UCLA SCTC GIT 2.0 in relation to IVIG administration. (A) Schematic figures of the study design. Japanese version of the GIT score was analyzed on day 1 and day 7. IVIG 2 g/kg was administered over 5 days (day 2-6). **(B)** The total GIT score and subscales before and after IVIG administration. *P* values were calculated by Wilcoxon signed-rank test. * *P* < 0.05.

### Discussion

In the present study, Japanese version of the UCLA SCTC GIT 2.0 instrument demonstrated commendable internal consistency and good reliability (**Table 2**), comparable with its original version[9]. Additionally, Japanese version of the GIT score exhibited robust divergent validity demonstrated by significant association with F-scale (**Table 3**), suggesting its usefulness as a tool for evaluating GIT symptoms associated with SSc in real clinical settings. GIT symptoms receive less attention than other symptoms of SSc; GIT manifestations are not evaluated in composite measures of the disease such as the American College of Rheumatology Composite Response Index in Systemic Sclerosis.[30] The absence of significant correlations between GIT score outcomes and other clinical manifestations of SSc affirmed that GIT involvement in SSc stands as an independent factor (**Supplementary Table 1**), warranting separate evaluation.

When contrasted with the original study utilizing the English version,[9] several baseline differences in the study population were observed (**Table 1**). The Japanese version assessment was conducted on a smaller patient population (n=38 vs. 152); the patients were older (mean age = 65 vs. 51 years); and our evaluation indicated lower mean scores in all the subscales: reflux (0.25 vs. 0.69), distention/bloating (0.70 vs. 1.07), fecal soilage (0.13 vs. 0.30), diarrhea (0.49 vs. 0.56), social functioning (0.21 vs. 0.26), emotional well-being (0.17 vs. 0.49), constipation (0.36 vs. 0.43), and total GIT score (0.29 vs. 0.66). One explanation might be the higher proportion of patients already treated; most of our patients had already on proton pump inhibitors (89%). Alternatively, one could interpret our study as having enrolled individuals with SSc who had comparatively milder disease manifestations and lesser health impairments. This interpretation finds support in our assessment of HRQOL, revealing a mean SF-36 PCS and MCS score of 41.2 and 50.4, respectively, in contrast to the original study where these scores were 36.7 and 47.1, respectively. Moreover, our study featured a smaller proportion of patients with dcSSc (22% vs. 55%), a factor associated with severe gastrointestinal involvement in SSc.[31]

An advantage of this study is the multi-dimensional immunophenotyping conducted, which encompassed assessments of serum cytokine levels and autoantibody profiles, aligned with the GIT score outcomes. As a result, we found that the serum levels of TNF-α, IL-6, and VEGF were significantly correlated with the specific subclass of the GIT score (**Figure 1**). Elevation of serum levels compared to healthy controls and correlation with clinical manifestations of SSc have been reported regarding TNF-α,[32] IL-6,[33] and VEGF.[34] The involvement of IL-6 in the pathogenesis of SSc has been strongly suggested by a substantial body of experimental evidence.[35] This is further supported by the approval of tocilizumab, an anti-IL-6 receptor monoclonal antibody, for the treatment of SSc-ILD by the United States Food and Drug Administration.[36] Meanwhile, the therapeutic effectiveness of TNF-α inhibitors in treating SSc has not yet been conclusively demonstrated through randomized placebo-controlled trials, despite some promising findings from smaller observational studies.[37] It’s worth noting that experimental studies involving biopsies of patients with GERD have revealed that cultured esophageal epithelial cells, fibroblasts, and muscle cells primarily produce IL-6, rather than TNF-α.[38] This finding opens the possibility of exploring the response of GERD symptoms in SSc patients to anti-IL-6 therapies, assessed using the GIT score, as a compelling avenue for future research investigations.

The primary highlight of this study lies in its ability to demonstrate the sensitivity of the GIT score through the improvement observed in the GIT score before and after IVIG administration (**Figure 2**). In a prior publication, we presented evidence of rapid alleviation of subjective symptoms and imaging findings of SSc-related GIT symptoms such as intestinal pseudo-obstruction, and moreover, weight recovery and weaning from total parenteral nutrition following regular monthly IVIG treatments in patients with SSc-myositis overlap.[7] Our current study reaffirmed the immediate effectiveness of IVIG, as reflected in the improvement of the GIT score. These finding underscores the utility of the GIT score as a tool for evaluating the effectiveness of IVIG in Japanese SSc patients within real-world clinical settings and, prospectively, in forthcoming clinical trials.

Our study has several limitations. Initially, it is important to note that the sample size in our study was relatively modest. This limitation could potentially explain our inability to detect any associations between autoantibody profiles and the GIT scores (**Supplementary Table 2**), even though certain autoantibodies, such as anti-U3-RNP Ab,[39] which are recognized for their association with GI involvement in SSc. Furthermore, it is worth acknowledging that our evaluation of GIT scores before and after IVIG administration followed a retrospective design. Consequently, the potential for biases cannot be entirely ruled out, although we made efforts to minimize them by sequentially enrolling cases. Additionally, it posed challenges to definitively differentiate the impact of SSc from myositis on GIT symptoms, as our assessment was limited to patients with SSc-myositis overlap. To comprehensively address the efficacy and safety of IVIG in managing SSc-related GIT symptoms, future studies should aim for a more rigorous investigation, ideally in a prospective, multicenter, randomized, and placebo-controlled setup.

## Contributor-ship

Conceptualization: KMM, AY

Methodology: KMM

Investigation: KMM, ES

Clinical data acquisition: KMM, YA, MK, MM, YN, HK, TH, AK, TF

Project administration: KMM

Supervision: AY, SS

Writing – original draft: KMM

Writing – review & editing: AY, SS

## Data Availability

All data produced in the present study are available upon reasonable request to the authors.

## Acknowledgements

We honor and appreciate Prof. Dinesh Khanna for developing and providing us with the original version of the UCLA SCTC GIT 2.0. We thank Ms. Maiko Enomoto and her colleagues for technical assistance and secretary work. We express our gratitude to Ms. Teruko Tani and Ms. Mayumi Odagaki for their assistance in collecting clinical information.

## Ethical approval information

This study was approved by The University of Tokyo Ethical Committee (Approval number 0695).

## Data sharing statement

The data analyzed during the current study are available from the corresponding author on reasonable request.

---

**Supplementary Figure 1.** Autoantibody profiling. **(A)** Schematic figures of A-Cube. Synthesized antigens were displayed on glass slides in an array format. The arrays were treated with patients’ serum, followed by application of fluorescence-labeled secondary antibodies specific to human IgG. The serum levels of each autoantibody were determined through analysis of the resulting fluorescence image. GSH: glutathione. **(B)** Correlational heatmap between each autoantibody. r: Spearman’s rho. The size of each circle represents the *P* value. **(C)** The UpSet plot of autoantibodies.

