## Supplementary Data 1 for "Reliability, validity, and sensitivity of Japanese version of the UCLA Scleroderma Clinical Trial Consortium Gastrointestinal Tract Instrument: application to efficacy assessment of intravenous immunoglobulin administration"

### UCLA SCTC GIT 2.0 Questionnaire

The following questions ask about your digestive (gastrointestinal) symptoms over the past 7 days and how much they have affected your life. For all questions, please choose your answers from the options as indicated. If you are not sure how to answer a question, please select the closest possible answer.

**<u>In the past week,</u> please indicate the frequency (number of days) that the following occurred.**

| | None | 1-2 days | 3-4 days | 5-7 days |
|---|---|---|---|---|
| Had difficulty swallowing solid foods. | ☐ | ☐ | ☐ | ☐ |
| Unpleasant sharp pain or burning sensation in the chest (heartburn) | ☐ | ☐ | ☐ | ☐ |
| Felt bitter or sour liquid coming up from the stomach to the mouth (gastric acid reflux) | ☐ | ☐ | ☐ | ☐ |
| Heartburn when eating "sour" foods like tomatoes and oranges | ☐ | ☐ | ☐ | ☐ |
| Vomited back up (vomited a small amount of food eaten or came up) | ☐ | ☐ | ☐ | ☐ |
| Slept with upper body elevated or back upright. | ☐ | ☐ | ☐ | ☐ |
| Had nausea/feeling sick | ☐ | ☐ | ☐ | ☐ |
| I vomited. | ☐ | ☐ | ☐ | ☐ |

**In the past week, please indicate the frequency (number of days) that the following occurred.**

| | None | 1-2 days | 3-4 days | 5-7 days |
|---|---|---|---|---|
| I had a feeling of fullness (gas or air in my stomach) | | | | |
| Sometimes the belly would bulge and sometimes it was necessary to loosen belts or unbutton pants and shirts | | | | |
| A little food filled me up. | | | | |
| Lots of farting and gas. | | | | |
| I didn't make it to the bathroom in time and soiled my underwear with stool. | | | | |
| Loose stools. | | | | |
| Stools were watery. | | | | |

**In the past week, please indicate how often your social life was interrupted by the following (e.g., visiting friends or relatives).**

| | None | 1-2 days | 3-4 days | 5-7 days |
|---|---|---|---|---|
| nausea | | | | |
| vomiting | | | | |
| gastralgia | | | | |
| diarrhea | | | | |
| Worries that you might accidentally soil your underwear | | | | |
| sensation of fullness | | | | |

**<u>In the past week,</u> please indicate the frequency (number of days) that the following occurred.**

| | None | 1-2 days | 3-4 days | 5-7 days |
|---|---|---|---|---|
| Felt concerned or anxious about gut issues | ☐ | ☐ | ☐ | ☐ |
| Embarrassment due to bowel symptoms. | ☐ | ☐ | ☐ | ☐ |
| Intestinal symptoms caused sexual problems and problems in the relationship with the partner | ☐ | ☐ | ☐ | ☐ |
| I was afraid I might not be able to find a restroom. | ☐ | ☐ | ☐ | ☐ |
| Mental depression or weakness due to intestinal symptoms | ☐ | ☐ | ☐ | ☐ |
| Avoided or postponed travel due to intestinal symptoms | ☐ | ☐ | ☐ | ☐ |
| Anger and irritability due to intestinal symptoms. | ☐ | ☐ | ☐ | ☐ |
| Sleep interrupted due to intestinal symptoms. | ☐ | ☐ | ☐ | ☐ |
| Felt bowel symptoms worsened due to "stress" or emotional disturbances | ☐ | ☐ | ☐ | ☐ |

**Over the past week,** have you noticed that your stools have become

| | No | Yes |
|---|---|---|
| It's getting hard. | ☐ | ☐ |

**In the past week,** please indicate how often the following occurred

| | None | 1-2 days | 3-4 days | 5-7 days |
|---|---|---|---|---|
| Constipation or failure to defecate | ☐ | ☐ | ☐ | ☐ |
| Stools were hard. | ☐ | ☐ | ☐ | ☐ |
| Pain during defecation. | ☐ | ☐ | ☐ | ☐ |
