## Supplementary figures and images for "Reliability, validity, and sensitivity of Japanese version of the UCLA Scleroderma Clinical Trial Consortium Gastrointestinal Tract Instrument: application to efficacy assessment of intravenous immunoglobulin administration"

### Supplementary Figure 1

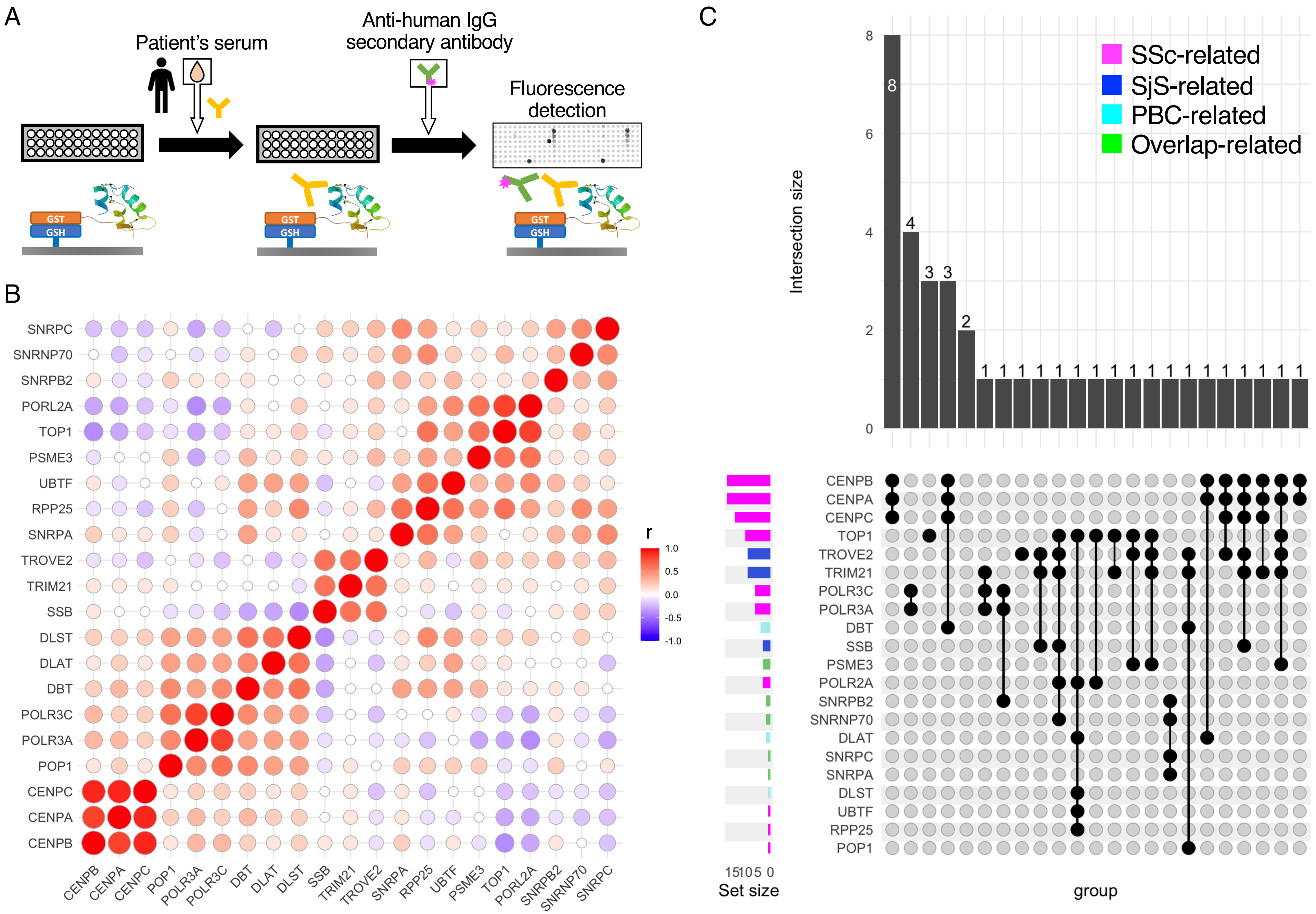


Matsuda KM et al.
Supplementary Figure 1
